# A novel hypothesis-generating approach for detecting phenotypic associations using epigenetic data

**DOI:** 10.1101/2024.01.15.24301219

**Authors:** Florence Z. Martin, Kayleigh E. Easey, Laura D. Howe, Abigail Fraser, Deborah A. Lawlor, Caroline L. Relton, Gemma C. Sharp

**Affiliations:** MRC Integrative Epidemiology Unit the University of Bristol, Bristol, UK; School of Psychological Science, University of Bristol, Bristol, UK; London School of Hygiene and Tropical Medicine, London, UK; Department of Psychology, University of Exeter, Exeter, UK

**Keywords:** hypothesis-generating, hypothesis-testing, epigenome-wide association study, dysmenorrhea, heavy menstrual bleeding, ALSPAC, ARIES

## Abstract

**Background:** Hypotheses about what phenotypes to include in causal analyses (that in turn can have clinical and policy implications) can be guided by hypothesis-free approaches, leveraging the epigenome for example. Materials & methods: Minimally adjusted epigenome-wide association studies (EWAS) using ALSPAC data were performed for example conditions, dysmenorrhea and heavy menstrual bleeding (HMB). Differentially methylated CpGs were searched in the EWAS Catalog and associated traits identified. Traits were compared between those with and without the example conditions in ALSPAC. Results: Seven CpG sites were associated with dysmenorrhea and two with HMB. Smoking and adverse childhood experience score were associated with both conditions in the hypothesis-testing phase. Conclusion: Hypothesis-generating EWAS can help identify associations for future analyses.

**Plain language summary:** To make a positive impact on policy and clinical practice, it is important that epidemiologists, those who study population health, can identify characteristics that might increase the risk of medical conditions. However, it can be difficult to know which associations should be investigated and decisions can often be biased by pre-formed opinions about what is relevant. In this study, we wanted to look for potential risk factors for dysmenorrhea (painful periods) and heavy menstrual bleeding (HMB) using a hypothesis-free approach (in other words, minimal adjustment for potential confounders), leveraging epigenetic data from a sub-sample of the Avon Longitudinal Study of Parents and Children (ALSPAC) and generating hypotheses about associations, then testing these hypotheses in the wider ALSPAC cohort. This meant looking for differentially methylated CpGs between those with and without the conditions of interest using an epigenome-wide association study (EWAS), seeing which phenotypes were associated with the CpGs in the EWAS Catalog, and testing these hypotheses in the ALSPAC cohort using measurements of each phenotype. For dysmenorrhea, we found seven differentially methylated CpGs and for HMB, we found two. These CpGs were associated with several phenotypes, which we could proxy in the wider ALSPAC cohort, creating hypotheses we tested using regression analyses. In the hypothesis-testing phase, we found that smoking and adverse childhood experience score were associated with dysmenorrhea and HMB. With this under-utilised approach, we can identify phenotypes that may be risk factors for under-studied conditions, that can be explored in other cohorts using analyses that can assess causality.

**Tweetable abstract:** Leveraging EWAS data can help identify novel potential risk factors for understudied conditions such as dysmenorrhea and heavy menstrual bleeding for future examination in causally motivated analyses: a proof-of-concept study in the Children of the 90’s cohort (ALSPAC)

## Introduction

Epigenome-wide association studies (EWAS) have been widely used in epidemiology over the last decade to explore biomarkers and aetiologies of health traits and disease (1). This explosion in use is due to both the volume and nature of the epigenetic data available to researchers, thanks to advances in bead-based microarray technology to measure levels of DNA methylation (DNAm) at individual CpG sites (2). The epigenome is a dynamic system of mitotically heritable markers that can control gene expression without changing the underlying genetic sequence; it can be altered by environmental exposures associated with a multitude of phenotypes (3). Traditional DNAm-based EWAS analyses aim to identify phenotype-CpG associations, under the assumption that these associations may either be causal (i.e., the CpG causes the phenotype or vice versa) or represent confounding in a way that could still be useful for indicating (historical) exposures or predicting future outcomes (4) (e.g., methylation at *AHRR* can be used to identify current and former smokers, and is predictive of lung cancer, even in the absence of causal mediation (5)). DNAm at specific CpGs can be thought of as phenotypes, because their variation will have both a genetic and an environmental basis (6). This feature means that, even in the absence of any causal epigenetic relationship, DNAm data may be useful for identifying (potentially causal) associations between other, non-epigenetic phenotypes, which can be followed by more causally motivated studies. This kind of hypothesis generation is particularly useful for under-researched phenotypes and conditions where limited previous literature is available to guide hypothesis-driven approaches.

Menstrual health is one such example of an under-researched area. For reasons related to entrenched gender inequalities and menstrual stigma (7), there are still knowledge gaps around the risk factors and consequences of experiencing problematic menstrual symptoms like menstrual pain (dysmenorrhea) and heavy or prolonged menstrual bleeding (HMB), both of which are difficult to quantify and diagnose. However, these are common symptoms that affect a large proportion of the female population throughout the life course that may impact on day-to-day life (8); these symptoms can also be considered important indicators of other domains of health and wellbeing (9). The prevalence of these in adolescence is estimated to be between 43% to 93% for dysmenorrhea (10) and 37% for HMB (11). Behaviours such as smoking and characteristics such as body mass index (BMI) have been linked inconclusively to dysmenorrhea, with a number – but not all – of studies reporting associations (12-15). The picture is even less clear for HMB outside comorbidities such as ovulatory dysfunction and coagulation disorders (16), with potential links to high body mass index, smoking, and alcohol consumption (17).

In this study, we aimed to use a minimally adjusted EWAS to leverage confounding and identify phenotypes that may be associated with dysmenorrhea and HMB as example conditions, where associations could be tested in the wider cohort data in a later phase. In this paper, we use the term phenotype to refer to any non-genetic characteristic that might be a potential risk factor. The approach, in short, involved: 1) identifying condition-CpG associations by running EWAS in the Avon Longitudinal Study of Parents and Children (ALSPAC) among those (G1) adolescents with epigenetic data; 2) looking up identified CpGs in an online repository of published EWAS results (EWAS Catalog (18)) to identify phenotypes associated with those CpGs (i.e., generating hypotheses about phenotypic associations); and 3) testing those hypotheses in the full ALSPAC cohort to explore associations between our conditions (dysmenorrhea and HMB) and the EWAS Catalog phenotypes (i.e., testing hypotheses). The two conditions under investigation here are understudied and prevalent, with little known about modifiable risk factors, thus making them useful case studies for proposed utility of the hypothesis-generating EWAS approach.

## Materials & methods

### Study population

ALSPAC is a longitudinal birth cohort study that recruited pregnant women between 1990 and 1992 in the previous area of Avon, in the Southwest of England (19). The initial sample consisted of 14,541 pregnancies enrolled in the first phase; a further 913 pregnancies were added at subsequent recruitment phases, leading to 14,901 index children remaining in the cohort after one year (19). A subset of 1,018 ALSPAC mother-child pairs, selected based on the availability of DNA samples, was included in the Accessible Resource for Integrative Epidemiologic Studies (ARIES) study, which generated DNA methylation data at three timepoints: birth, childhood, and adolescence (20).

The initial eligibility criteria for this study were female index children (G1 participants) who had responded to at least one of the nine “G1 puberty” questionnaires sent between the ages of 8 and 17 years, specifically the section pertaining to menstruation (whether it had started, whether there were issues associated with it, etc.). Among those who fulfilled the initial eligibility criteria, we then identified those who had participated in ARIES and had methylation data (described below). The exclusions are summarised in the flow diagram (Figure 1).

**FIGURE 1.**
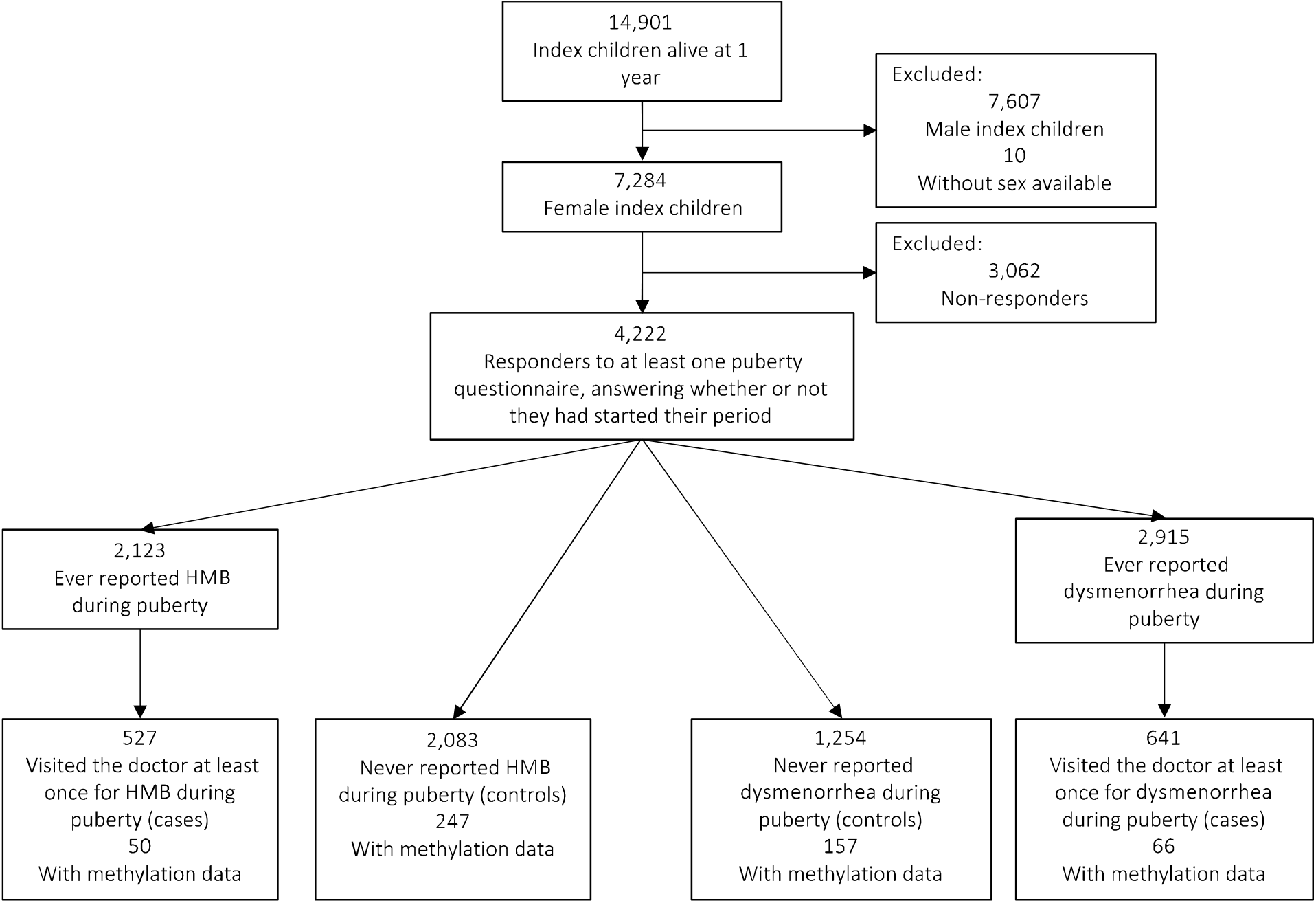
FLOW OF PARTICIPANTS THROUGH THE STUDY.

Ethical approval for the study was obtained from the ALSPAC Ethics and Law Committee and the Local Research Ethics Committee. Informed consent for the use of data collected via questionnaires and clinics was obtained from participants following the recommendations of the ALSPAC Ethics and Law Committee at the time. Consent for biological samples has been collected in accordance with the Human Tissue Act 2004. For data collection from participants at 22 years old and onwards, study data were collected and managed using REDCap (Research Electronic Data Capture) electronic data capture tools hosted at the University of Bristol (21). The ALSPAC study website contains details of all the data that is available through a fully searchable data dictionary and variable search tool (http://www.bristol.ac.uk/alspac/researchers/our-data/) (22).

### Definition of menstrual symptoms

In each of the nine G1 puberty questionnaires, the participants’ caregiver (age 8–13) or participant (age 14 and over) was asked “Have you/your daughter ever had any of the following symptoms associated with your/her period: Severe cramps?” and “Heavy or prolonged bleeding?”. Answering “Yes” to each question prompted a question asking if a doctor was contacted for the symptom. Other than at age 15 for cramps, we did not have self-reported severity for either symptom, so for the purpose of the current study, we used “ever having visited the doctor” to define dysmenorrhea or HMB. The menstrual health-related data in ALSPAC is summarised in detail elsewhere (23).

### Identifying novel traits associated with menstrual symptoms

The approach is detailed in Figure 2 and below.

**FIGURE 2.**
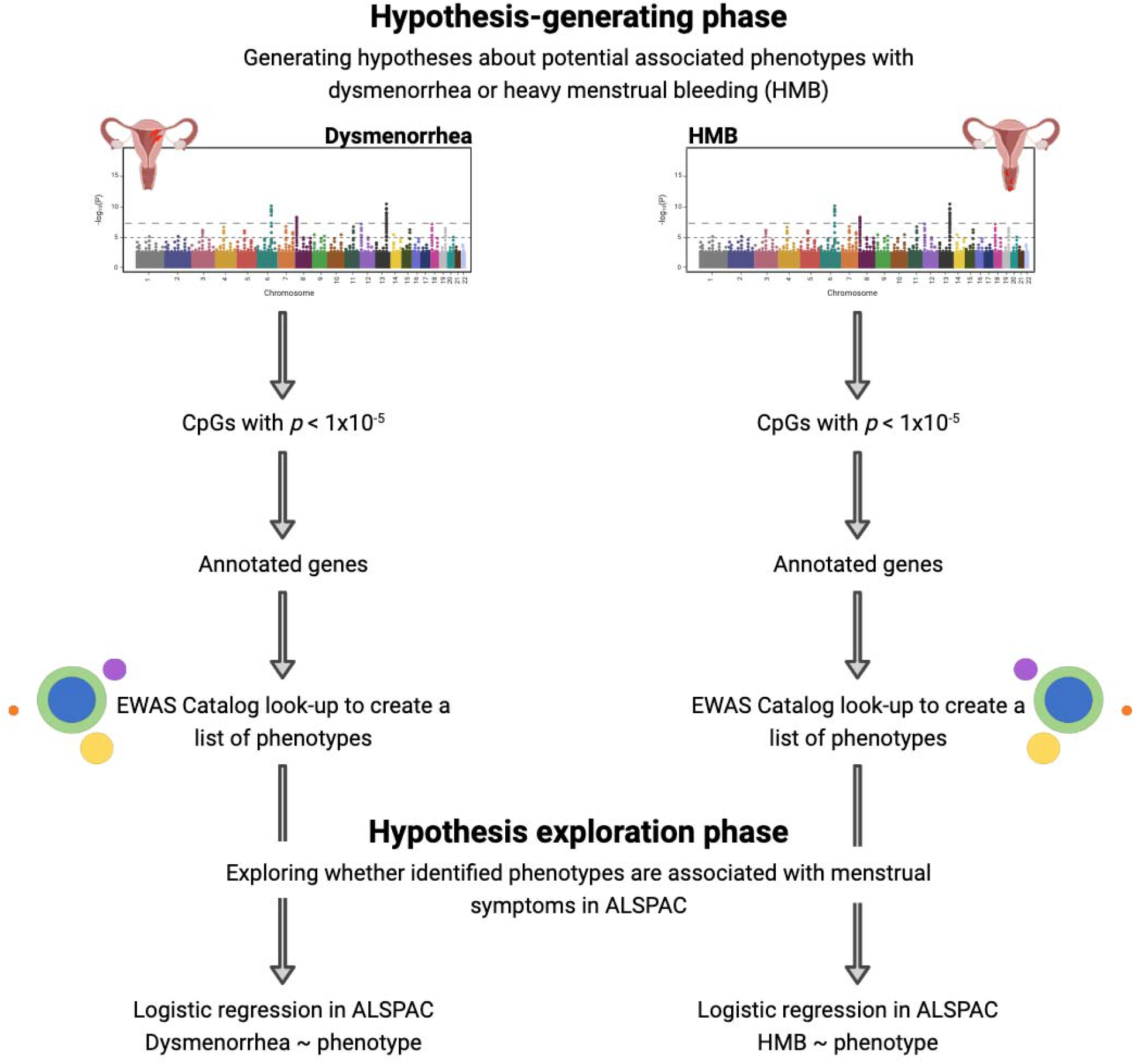
OUR METHODOLOGY FOR IDENTIFYING NOVEL PHENOTYPES ASSOCIATED WITH DYSMENORRHEA AND HMB IN ADOLESCENTS USING A HYPOTHESIS-GENERATING EPIGENOME-WIDE ASSOCIATION STUDY APPROACH. CREATED WITH BIORENDER.COM.

### Hypothesis-generating phase

Generation and preparation of the DNA methylation data for the subsample of ALSPAC known as ARIES is described in detail in the ARIES Data Resource Profile (20). Briefly, DNA methylation in peripheral blood samples obtained from 1,018 adolescents (aged 15 or 17 years old) was measured using the Illumina Infinium® HumanMethylation450K BeadChip assay (20). This array contains probes that can measure the methylation of over 450,000 CpGs located throughout the human genome, driven by DNA bisulfite conversion (24). The level of methylation is given as a beta-value (*b*), ranging from not methylated (*b*=0) to fully methylated (*b*=1) (25).

Data obtained from the array were pre-processed and quality controlled using the R package meffil (26). We removed 11,648 probes that mapped to either the X or Y chromosomes, 901 control probes, and 3,853 probes that had a high detection *P*-value. Following the removal of these probes, samples with outlying methylation values were identified using the Tukey method (outside the 25^th^ and 75^th^ percentiles ±3 times the interquartile range) (27) and removed for individual probes.

### Epigenome-wide association studies – Hypothesis-generating phase

This was explored using a cross-sectional design within ALSPAC (i.e., both DNA methylation and menstrual symptoms were measured at ages that overlapped). Association between dysmenorrhea/HMB case/control status and variation in methylation was assessed using linear regression. Case status (dysmenorrhea or HMB any time during adolescence) was treated as the exposure in the EWAS, with methylation included as the outcome, since our aim was not to identify causal associations. Models were thus purposefully simple: we adjusted only for age at methylation measurement (given that the adolescent peripheral blood samples were collected at either age 15 or 17) and surrogate variables generated to capture technical batch effects. Surrogate variables (SV) were generated using surrogate variable analysis (SVA) and the number of surrogate variables to generate was estimated as part of the SVA pipeline (*nSV*=24 and *nSV*=33 for dysmenorrhea and HMB EWAS, respectively). All analyses were performed using the R package meffil (which draws on the R package isva to generate surrogate variables) (26) using R version 3.6.3. To enable the methylation data to capture variation in a wide range of other traits, we performed no further adjustment, e.g., for cell counts or hormonal contraception.

### EWAS Catalog look up

From each EWAS (of dysmenorrhea and HMB), we selected differentially methylated CpGs with a false discovery rate (FDR)-adjusted *P*-value<1×10^-5^ and performed a look-up of these CpGs, or the genes they mapped to, in the EWAS Catalog (18). The EWAS Catalog is a repository of phenotype-CpG associations identified through published EWAS. We created a list of phenotypes associated with these CpGs; although pleiotropy was present for some CpGs, an enrichment analysis was not deemed necessary due to the hypothesis-generating nature of the analysis.

### Hypothesis-testing phase in full ALSPAC sample

We used the ALSPAC data dictionary to find data on corresponding CpG-associated phenotypes in ALSPAC, assessed during gestation (for prenatal exposures) or pre-puberty (for childhood traits). Pre-puberty measurements of traits were chosen to determine whether the identified phenotype preceded the onset of these symptoms, and thus may be a candidate for future testing in causal analyses in other datasets as potential risk factors for either menstrual symptom. Further details of this process are described in the supplementary material. To explore the phenotypes identified in the look up phase, we performed logistic regressions with each identified phenotype included in turn as the exposure and the conditions (HMB or dysmenorrhea) in ALSPAC adolescents as the outcome, with and without adjustment for socioeconomic position (SEP) and age at menarche (AAM). Continuous variables were converted to standardised z-scores before running these analyses to enable comparison of effect estimates for variables on different scales. Phenotypes where numbers of cases and/or controls contained fewer than five participants were omitted from the analysis due to inadequate power.

To test previously identified associations between phenotypes such as gynaecological and endocrine disorders, socioeconomic position, oral contraception, and age at menarche and menstrual symptoms, we also included these alongside the series of logistic regressions of novel associations.

### Sensitivity analyses

We ran the EWAS analysis for each symptom again removing cases of thyroid problems (self-reported at 17 years), polycystic ovary syndrome (PCOS) and endometriosis (self-reported at 22 years) to limit the effect these conditions might have had on the findings. We also replicated the hypothesis-testing phase among participants who had reported the symptom during puberty but had not visited the doctor, to identify whether any characteristics were associated with a less “severe” presentation of dysmenorrhea or HMB. We investigated use of oral contraception in the hypothesis-testing phase rather than excluding them given the high prevalence of use among adolescents.

## Results

Of the original ALSPAC cohort, 7,284 participants were female (49% of the children who were alive at 1 year old) and 4,222 of these participants responded to at least one of the questionnaires sent out during puberty, stating whether they had started their period. Of these, 487 individuals had DNA methylation data at adolescence in ARIES (Figure 1) and were included in the hypothesis-generating phase. In the hypothesis-testing phase, we performed complete case analyses so the denominator differed by analysis depending on the missing data for each phenotype (supplementary tables 1 and 2).

### Hypothesis-generating phase

#### Dysmenorrhea

We identified seven differentially methylated CpG sites (FDR-adjusted *P*<1×10^-5^) (Table 1) in adolescents who suffered from severe dysmenorrhea compared with those who did not. DNA methylation at these CpG sites was associated with 31 phenotypes in the EWAS Catalog look-up.

**TABLE 1.**
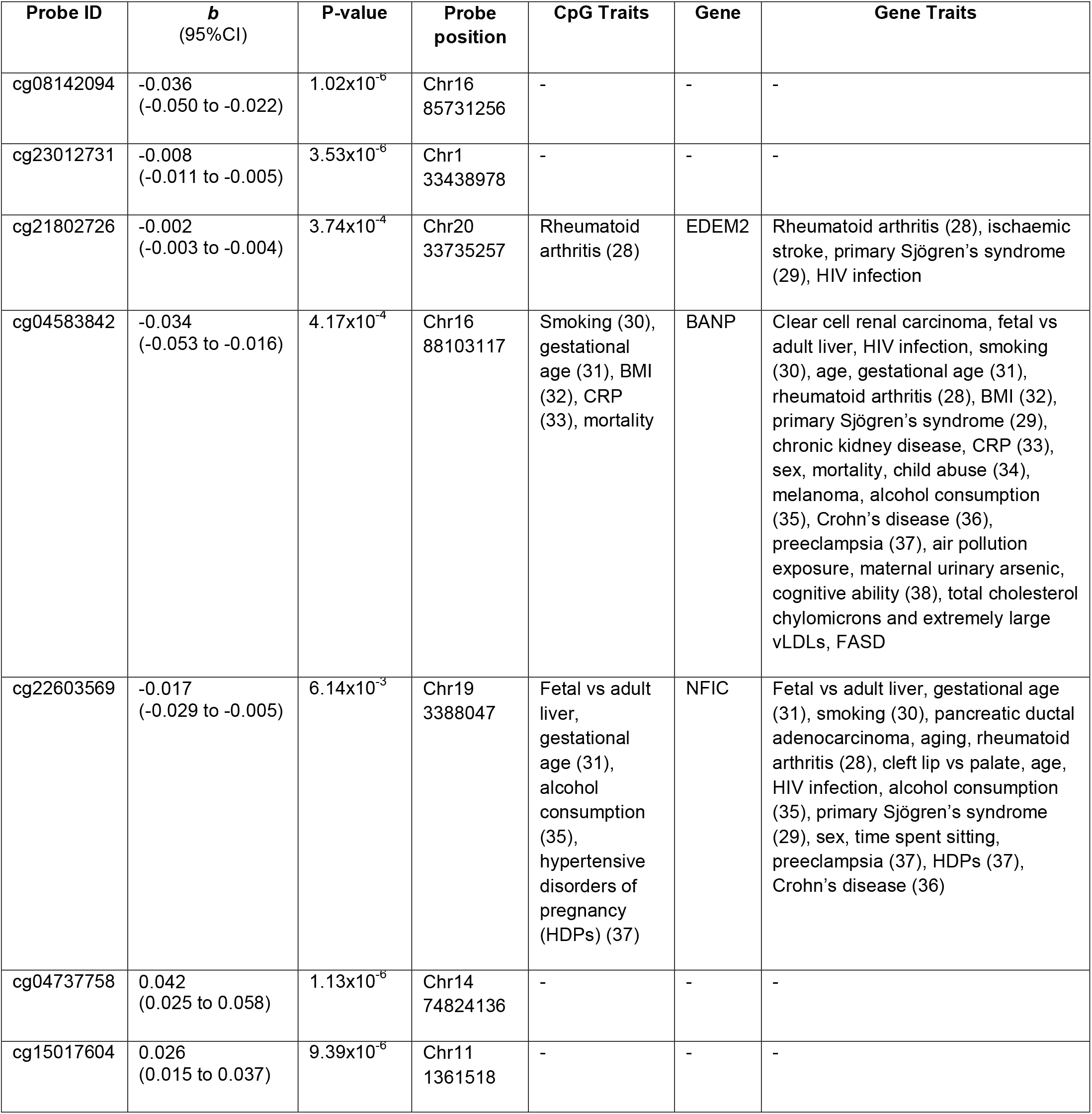
DIFFERENTIALLY METHYLATED CPG SITES IDENTIFIED IN THE HYPOTHESIS-GENERATING EWAS OF DYSMENORRHEA. PROBE ID DENOTES THE DIFFERENTIALLY METHYLATED CPG SITE. B REPRESENTS THE EXTENT OF METHYLATION, WITH FDR-CORRECTED RESPECTIVE P-VALUES. PROBE POSITION REPRESENTS THE CHROMOSOMAL LOCATION OF THE CPG SITE, WHILE CPG TRAITS DENOTES TRAITS THAT HAVE BEEN IDENTIFIED TO BE ASSOCIATED WITH THAT CPG SITE. THE GENE COLUMN SHOWS IN WHICH GENE THE CPG SITE IN QUESTION RESIDES, WITH GENE TRAITS REPRESENTING TRAITS THAT HAVE BEEN SHOWN TO BE ASSOCIATED WITH CPG SITES LOCATED WITHIN THAT GENE.

#### HMB

In the EWAS of HMB, we identified two differentially methylated CpG sites with an FDR-adjusted *P*<1×10^-5^ (Table 2). When we performed a look-up of these CpGs, as well as the genes they mapped to, in the EWAS Catalog, we identified 10 associated phenotypes.

**TABLE 2.** DIFFERENTIALLY METHYLATED CPG SITES IDENTIFIED IN THE HYPOTHESIS-GENERATING EWAS OF HMB. PROBE ID DENOTES THE DIFFERENTIALLY METHYLATED CPG SITE. β REPRESENTS THE EXTENT OF METHYLATION, WITH FDR-CORRECTED RESPECTIVE P-VALUES. PROBE POSITION REPRESENTS THE CHROMOSOMAL LOCATION OF THE CPG SITE, WHILE CPG TRAITS DENOTES TRAITS THAT HAVE BEEN IDENTIFIED TO BE ASSOCIATED WITH THAT CPG SITE. THE GENE COLUMN SHOWS IN WHICH GENE THE CPG SITE IN QUESTION RESIDES, WITH GENE TRAITS REPRESENTING TRAITS THAT HAVE BEEN SHOWN TO BE ASSOCIATED WITH CPG SITES LOCATED WITHIN THAT GENE.

| Probe ID | $\beta$<br>(95%CI) | p-value | Probe<br>position | CpG Traits | Gene | Gene Traits |
| --- | --- | --- | --- | --- | --- | --- |
| cg24196053 | -0.010<br>(-0.014 to -0.006) | $1.50 \times 10^{-5}$ | Chr4<br>1736433 | - | TACC3 | Clear cell renal carcinoma, gestational age (31), HIV infection, Crohn's disease (36), Alzheimer's disease, preeclampsia (37), total cholesterol |
| cg11465939 | -0.029<br>(-0.042 to -0.015) | $6.04 \times 10^{-5}$ | Chr22<br>23923462 | Fetal vs adult liver, smoking (30), gestational age (31) | IGLL1 | Clear cell renal carcinoma, fetal vs adult liver, gestational age (31), HIV infection, primary Sjögren's syndrome (29), smoking (30) |

### Hypothesis-testing phase

From the list of phenotypes that were associated with differentially methylated CpGs in the hypothesis-generating phase (Table 1 and Table 2), we derived the following variables from ALSPAC: maternal educational attainment, maternal smoking and alcohol consumption during pregnancy, maternal body mass index (BMI), maternal hypertensive disorders of pregnancy (HDP), maternal preeclampsia, participant gestational age at delivery, participant BMI, cotinine and cholesterol levels at age 7, participant non-word repetition score (measure of cognition) at age 8, participant C-reactive protein (CRP) at age 9, participant cigarette and alcohol use at age 13 and participant adverse childhood experience (ACE) score by 16 years old (39). Of these, participant smoking and BMI had been identified *a priori* as associated with both menstrual symptoms. We then compared these variables for cases and controls for each condition in the wider ALSPAC cohort (*n*=4,222) Other characteristics such as kidney disease, primary Sjögren’s syndrome, melanoma, Crohn’s disease, and rheumatoid arthritis were available, but cases and/or controls for each symptom contained fewer than five participants so were omitted from the analysis as described in the methods.

We found that, compared to unexposed participants, participants who had been exposed to smoke prenatally and who had smoked or drunk alcohol by age 13 were more likely to have dysmenorrhea in the unadjusted models (Figure 3). Higher BMI, CRP, cotinine, and ACE score was also associated with an increased risk of dysmenorrhea (Figure 4). Following adjustment for SEP and AAM, these effects attenuated, except for smoking at 13 years and ACE score at 16 years (aOR 1.61 95%CI 1.11–2.33 and aOR 1.30 95%CI 1.11–1.53, respectively (supplementary tables 3 and 4).

**FIGURE 3.**
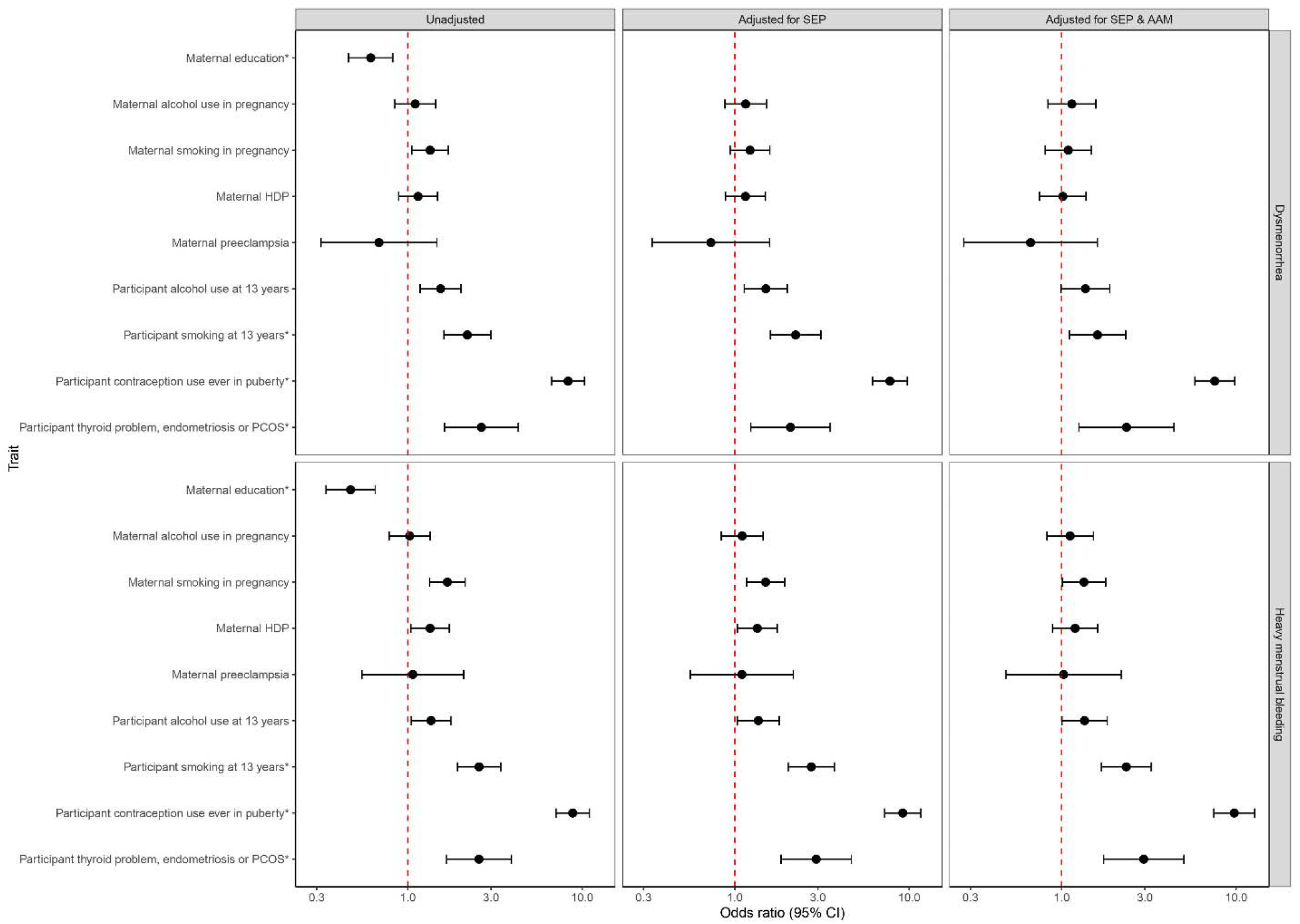
COEFFICIENT PLOT REPRESENTING BINARY PHENOTYPES ASSOCIATED WITH DYSMENORRHEA AND HMB IN THE HYPOTHESIS TESTING PHASE. * IDENTIFIED AS AN ASSOCIATED TRAIT A PRIORI

**FIGURE 4.**
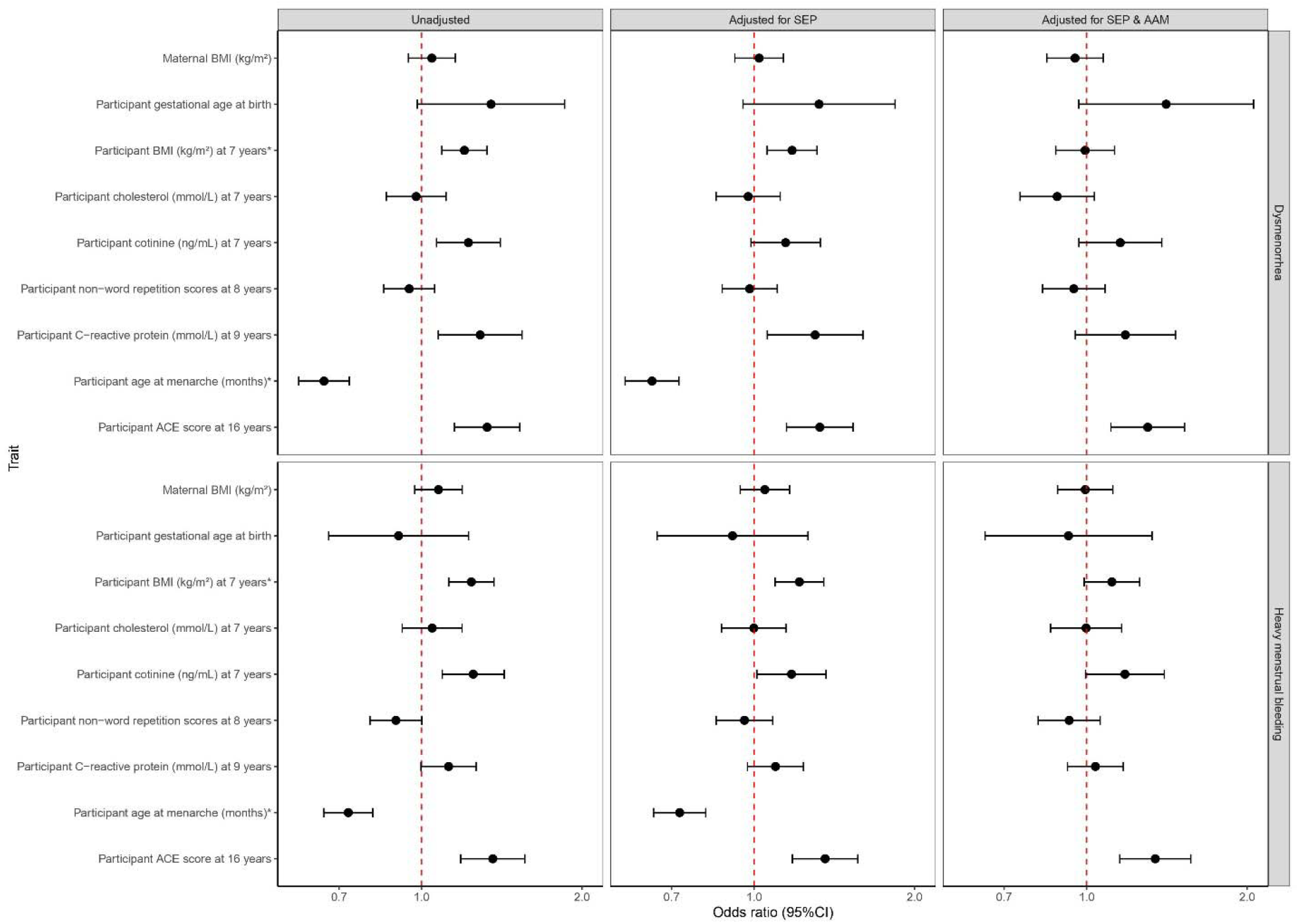
COEFFICIENT PLOT REPRESENTING CONTINUOUS PHENOTYPES ASSOCIATED WITH BEING A DYSMENORRHEA OR HMB CASE IN THE HYPOTHESIS TESTING PHASE. * IDENTIFIED AS AN ASSOCIATED TRAIT A PRIORI

In the unadjusted HMB models, participants exposed to smoke and maternal HDP prenatally were at a higher risk of developing HMB during puberty; smoking and drinking alcohol by the age of 13 was also associated with an increased risk of HMB (Figure 3). Higher BMI, cotinine, and ACE score was associated with an increased risk of HMB in the unadjusted models (Figure 4). Following adjustment for SEP and AAM, these effects attenuated, except for smoking at 13 years and ACE score at 16 years (aOR 2.35 95%CI 1.69–3.26 and aOR 1.35 95%CI 1.15–1.57, respectively) (supplementary tables 5 and 6).

Additionally, although all confidence intervals crossed the null, dysmenorrhea (but not HMB) was consistently associated with higher gestational age at delivery with relatively large effect estimates (aOR 1.41 95%CI 0.97–2.06) (Figure 4).

### Sensitivity analysis

In the sensitivity analysis, where we ran the same EWAS analysis for each symptom with cases of thyroid problems at 17 years and PCOS or endometriosis at 22 years removed, four hits from the primary analysis persisted for dysmenorrhea (supplementary table 7) and one hit persisted for HMB (supplementary table 8). The effect estimates in the sensitivity analysis all followed the same direction as the estimates from the primary analysis, and on the most part strengthened, except for cg04737758 which attenuated towards the null. We then ran the hypothesis-testing phase in the cohort with these cases removed. The direction of the associations with all traits didn’t change and the conclusions made from the primary analysis didn’t change (supplementary figures 1 and 2).

When we performed the hypothesis-testing phase in less severe cases of each symptom, most associations attenuated towards the null. Associations that were stronger in the sensitivity analysis of less severe cases (exposure to maternal preeclampsia and cotinine at age 7 for HMB and gestational age at birth for dysmenorrhea) were in the same direction as the primary analysis (supplementary figures 3 and 4).

## Discussion

In this study, we corroborated previously identified associations (i.e., BMI and smoking) and generated new hypotheses about phenotypes (i.e., adverse childhood experiences and alcohol consumption) that may be contributing to the development of adolescent dysmenorrhea and HMB. These hypotheses warrant further investigation in a causally motivated framework, as what we present here is purely associational. We were able to replicate previously reported associations with own smoking (12, 40) and higher BMI (15, 41) with both menstrual symptoms, guided by methylation markers. We believe that this is the first study to present evidence that that early life experiences such as ACEs and prenatal exposures such as maternal smoking are associated with these conditions. The identification of both previously identified associations, as well as novel condition-phenotype relationships, suggests that the use of a hypothesis-generating EWAS approach may be useful to identify associations for future causal inference work.

### Previous literature

EWAS investigate differentially CpGs in relation to a single phenotype and generates hypotheses, as the associations they identify between a trait of interest and differentially methylated CpGs may either be causal (i.e., the CpG/gene causes the trait), represent a historical exposure (i.e., flags someone as a smoker, for example), or highlight confounding. All these potential explanations for associations are useful when thinking about disease aetiology and future analyses (4). Their findings can focus subsequent causal analyses and can be implemented in scenarios where epigenetic data, as well as rich phenotypic data, are available. Genome-wide association studies (GWAS) are useful in such scenarios but only focus on genetics, whereas EWAS allows us to leverage confounding by incorporating exposures throughout the early life that might be associated with later life conditions. In the context of exposome-wide association studies (ExWAS), EWAS has been employed to reduce exposome dimension and make efficiency gains (42), reflecting the intention in this present study but here, to improve efficiency in subsequent observational, non-WAS analyses.

In order to test the combination of minimally adjusted EWAS with the hypothesis-testing phase for identifying associations, we wanted to use example conditions where few associations have been previously confirmed, so that there would be scope to identify novel traits. ALSPAC has repeat measures of the presence of dysmenorrhea and HMB throughout adolescence. In the literature, known causes and risk factors (outside diagnosed gynaecological problems) are scant, with some evidence suggesting smoking (40) and BMI (15, 41) are associated with these conditions.

### Novel findings

Having performed two EWAS, one for dysmenorrhea and the other for HMB, we identified seven and two differentially methylated CpG sites, respectively. The seven dysmenorrhea CpG hits were associated with 31 individual traits, including negative associations with smoking (previously identified) and alcohol consumption, child abuse, and preeclampsia (novel). The two HMB CpG hits were associated with ten individual traits including smoking (previously identified) and gestational age, total cholesterol, and preeclampsia (novel).

In the hypothesis-testing phase, we identified that smoking and alcohol consumption at age 13, as well as ACE score at age 16 were all associated with dysmenorrhea and HMB, including after adjustments for SEP and AAM. Although causal effects cannot be inferred from these analyses, they provide evidence that further investigation into these traits may be able to illuminate mechanisms by which they are associated with dysmenorrhea and HMB.

### Strengths and limitations

We present a potentially useful, epigenetic-based approach that can be implemented by leveraging confounding to identify phenotype-phenotype associations, provided there is sufficient access to epigenetic and phenotypic data, and carried out successfully even on a small scale. The small number of cases and controls limit our concerns that our findings may be a result of multiple testing. Given that the hypothesis-testing phase relies on a minimally adjusted EWAS analysis, a full complement of confounders is not required, thus participants are less frequently excluded for not having complete covariate data, which is particularly useful if case and control numbers are small. The G1 puberty questionnaires in ALSPAC were sent out multiple times allowing us several timepoints across adolescence within which to identify cases for our conditions. Despite cord blood methylation being available in ALSPAC, we chose to use adolescent DNA methylation because we wanted to identify exposures across the lifecourse to date that may be associated with the development of these conditions, additionally to genotypic differences.

A major limitation in this study was that, although the smaller numbers of tests being conducted provide reassurance against multiple testing (compared to a phenome-wide association study, for example), they are much smaller than those in other EWAS. However, prior work leveraging small number of participants in epigenetic analyses has driven hypotheses in other fields; for example, a small hypothesis-generating EWAS of paternal smoking identified offspring DNA methylation that might be associated with development (43) that was carried forward by another group investigating drivers of childhood autism (44). Also, despite our best efforts to maintain sensible temporality in the hypothesis-testing phase, sometimes we were not able to derive trait variables (exposure) that were definitively before the condition onset (outcome). For example, the ACE score variable used to explore the potential association between child abuse and the worked example conditions was a composite variable of ACEs up to the age of 16, given as a score (39). It is likely that some of the participants in the hypothesis-testing phase analysis of ACE score may have had ACEs that contributed to their score at age 15 or 16, where the onset of their dysmenorrhea was earlier in puberty. However, as we weren’t doing any causal analyses, merely attempting to identify associations between potential risk factors and menstrual conditions, temporality wasn’t of utmost importance to uphold. The findings from our hypothesis-testing phase of dysmenorrhea and HMB were only internally validated in the wider ALSPAC cohort, as opposed to in another cohort. It is important that future studies investigating the potential associations identified here do so in other independent cohorts where menstrual health data are available. The associations we observe in the hypothesis-testing phase may be completely mediated or modified by the much higher prevalence of hormonal contraception use in those with either condition; it is important to investigate the role of contraception for each association separately in future analyses. As Sawyer *et al*. point out, conditioning on hormonal contraception use might introduce collider bias if our phenotype of interest and condition are likely to influence contraception use (23). Finally, we are aware that the definition for severity for each condition (those who had visited a doctor for the symptom) may instead reflect socioeconomic, cultural, and personal factors for certain participants that will have influenced why they sought medical advice for their symptoms while others did not, which has been highlighted in other fields (45).

## Conclusions

We used an epigenome-led approach to generate hypotheses regarding potential risk factors, using dysmenorrhea and HMB as example phenotypes. The novel approach used here, leveraging both a hypothesis-generating and -testing phase, as well as confounding relationships, detected both known and novel associations between menstrual symptoms and environmental or physiological exposures. This novel approach could be added to the arsenal of exploratory analyses that drive hypotheses for future causal analyses in a range of understudied health problems, including menstrual health epidemiology.

## Data Availability

These data can be obtained from ALSPAC through the standard ALSPAC research proposal and data access policy (project number B3469)

http://www.bristol.ac.uk/alspac/researchers/access/

## Summary points

- DNA methylation can be explored in an epigenome-wide association study (EWAS) context to identify causal mechanisms or confounding relationships between exposures that can alter the epigenome and phenotypes of interest.
- In a minimally adjusted EWAS acting as hypothesis-generating, associations can be identified that represent either one of these relationships to be further explored in causally motivated analyses for conditions that are understudied.
- In the present study, we demonstrated the utility of a hypothesis-generating EWAS approach followed by a hypothesis-testing phase using logistic regression, investigating two understudied conditions as examples: dysmenorrhea (painful periods) and heavy menstrual bleeding (HMB).
- We used the Avon Longitudinal Study of Parents And Children (ALSPAC) to identify cases of adolescent dysmenorrhea and HMB; those with epigenetic data from ARIES were included in two hypothesis-generating EWAS where each condition served as the exposure.
- Having found seven differentially methylated CpGs for dysmenorrhea and two for HMB, we searched them in the EWAS Catalog and identified phenotypes associated with each of them.
- In the hypothesis-testing phase, we proxied the phenotypes found in the EWAS Catalog using variables from ALSPAC and included them in minimally adjusted logistic regression models where each condition served as the outcome.
- Using this approach, we found that smoking and alcohol use at age 13 was associated with dysmenorrhea and HMB; higher cotinine levels at age 7 was associated with HMB. Higher adverse childhood experience (ACE) score was associated with both conditions.
- Using this approach, we identified several potential targets of investigation for future research into risk factors for dysmenorrhea and HMB. Although temporality was not easily established in the present study and causality indeterminable, we leverage confounding to guide future causally motivated analyses in other cohorts with menstruation data.

## Reference annotations

Sharp GC, Relton CL. Epigenetics and noncommunicable diseases. Epigenomics. 2017;9(6):7 * of interest

An excellent and succinct editorial on the role of epigenetics in noncommunicable disease epidemiology.

Committee Opinion No. 651: Menstruation in Girls and Adolescents: Using the Menstrual Cycle as a Vital Sign. Obstetrics & Gynecology. 2015;126(6):e143-e6. * of interest

A recommendation highlighting the importance of menstrual monitoring from the start of menarche as part of a preventative approach for health issues in adulthood.

Schoep ME, Nieboer TE, van der Zanden M, Braat DDM, Nap AW. The impact of menstrual symptoms on everyday life: a survey among 42,879 women. Am J Obstet Gynecol. 2019;220(6):569 e1- e7. * of interest

Findings from a survey of over 40,000 women outlining the prevalence and burden of menstrual symptoms, like dysmenorrhea and HMB, on daily life.

Battram T, Yousefi P, Crawford G, Prince C, Sheikhali Babaei M, Sharp G, et al. The EWAS Catalog: a database of epigenome-wide association studies. Wellcome Open Res. 2022;7:41. * of interest

Data profile for the EWAS Catalog database that has collated CpG-trait associations from published EWAS.

Sawyer G, Howe LD, Fraser A, Clayton G, Lawlor DA, Sharp GC. Menstrual cycle features in mothers and daughters in the Avon Longitudinal Study of Parents and Children (ALSPAC). Wellcome Open Res. 2023;8:386. ** of considerable interest

Important data note summarising and describing all the menstrual health-related data in ALSPAC, some of which was used in the present study.

Morkve Knudsen GT, Rezwan FI, Johannessen A, Skulstad SM, Bertelsen RJ, Real FG, et al. Epigenome-wide association of father’s smoking with offspring DNA methylation: a hypothesis-generating study. Environ Epigenet. 2019;5(4):dvz023. ** of considerable interest

An example of the hypothesis-generating EWAS approach being used in a different setting: father’s smoking and offspring DNA methylation.

## Notes

### Competing Interest Statement

The authors have declared no competing interest.

### Funding Statement

The UK Medical Research Council and Wellcome (Grant number: 217065/Z/19/Z) and the University of Bristol provide core support for ALSPAC. This publication is the work of authors Florence Z. Martin, Dr Kayleigh E. Easey, Professor Caroline L. Relton, Professor Laura D. Howe, Professor Deborah A. Lawlor, Professor Abigail Fraser and Dr Gemma C. Sharp and Florence Z. Martin, Dr Kayleigh E. Easey, and Dr Gemma C. Sharp will act as guarantors for the contents of this paper. This research was funded in whole, or in part, by the Wellcome Trust (Grant number: 218495/Z/19/Z). For the purpose of Open Access, the author has applied a CC BY public copyright license to any Author Accepted Manuscript version arising from this submission. A comprehensive list of grants funding is available on the ALSPAC website (http://www.bristol.ac.uk/alspac/external/documents/grant-acknowledgements.pdf); Gemma C. Sharp is funded by a Medical Research Council New Investigator Research Grant (MR/S009310/1). This work is supported by the Menarche, Menstruation, Menopause and Mental Health (4M) consortium, established with grants from the GW4 (GW4-GF2-005; GW4-DA22-002).

### Author Declarations

Avon Longitudinal Study of Parents And Children (ALSPAC) Ethics and Law Committee of University of Bristol and Local Research Ethics Committee gave ethical approval for this work

