## supplementary material for "A novel hypothesis-generating approach for detecting phenotypic associations using epigenetic data"

### Replication of previously reported associations

In order to describe cases and controls at the age of 17, we compared characteristics that have been previously shown to be associated with dysmenorrhea or heavy menstrual bleeding (HMB). Characteristics included age at menarche (AAM), body mass index (BMI), hormonal oral contraception use, diagnosed endocrinological or gynaecological disorders (thyroid problems, polycystic ovary syndrome (PCOS) and endometriosis), smoking status, and socioeconomic position (SEP). We derived variables for these characteristics as follows. Age at menarche (AAM) was derived from the earliest report of menarche according to the puberty questionnaires, which were completed either by the adolescent themselves or their caregiver (depending on the age of the adolescent). Body mass index (BMI) was derived from height and weight measured at clinics at age 15 and 17, calculated as kg/m^2^; self-report BMI was also available at age 17 and was used for participants who did not have clinic measured BMI available. Each puberty questionnaire also asked adolescents if they had used oral contraception within the last 12 months; participants were classified as ever having used oral contraception during puberty if they had answered “yes” in any of the nine questionnaires. Thyroid problems were self-reported at age 17 (puberty questionnaire 9) and polycystic ovary syndrome (PCOS) and endometriosis were self-reported (participants answering “yes – self-diagnosed or “yes – doctor-diagnosed” were combined for the presence of either gynaecological disorder in a binary yes/no variable) at age 22 (1). We used cotinine measurements at age 17 to estimate smoking status, with a threshold of >10ng/mL used to indicate regular smoking (2). Highest maternal educational attainment was self-reported by the mother during pregnancy and used as a measure of participant SEP.

We summarised these characteristics by calculating the mean and standard deviation (continuous) or sample size and percentage (categorical) separately for dysmenorrhea and HMB cases and controls. Characteristics, both previously identified and novel, were not reported if either case or control groups contained fewer than five participants.

### ALSPAC variables

Following identification of further characteristics via our hypothesis-generating EWAS approach, we derived the following variables as early in puberty as possible, to determine whether associations preceded puberty and thus may be considered risk factors. Any alcohol and smoking exposure during pregnancy was derived from mother-completed questionnaires sent throughout gestation. Hypertensive disorders of pregnancy (HDP) and preeclampsia were abstracted from obstetric clinical records. Maternal pre-pregnancy BMI was calculated from self-reported height and weight at around 12 weeks’ gestation. Gestational age was calculated from mother’s last menstrual period (LMP); where this was uncertain or unavailable, the ultrasound assessment was used. Where neither of these were available, an obstetrician reviewed the notes and gave an estimate. BMI at age 7 was derived by height and weight taken at the Focus@7 clinic, and measurements of cotinine and cholesterol were taken at this clinic also. Non-word repetition scores (a short-term memory measure) was obtained at the Focus@8 clinic age 8, as this test captured cognitive score is the closest fashion as the EWAS that investigated this trait. C-reactive protein (CRP) was measured at age 9 at the Focus@9 clinic. At the Teen Focus 1 clinic, participants were asked whether they had ever tried smoking or drinking alcohol (mean age 12.8 years). Adverse childhood experience (ACE) score was derived from over 500 questions within ALSPAC referring to typical ACEs (forms of abuse, parental violence etc.) and a cumulative score generated; derivation of this variable is described in detail elsewhere (3).

**Supplementary Table 1** Number of each binary characteristic in cases and controls for each symptom identified in the hypothesis-generating EWAS and missing data in each variable

|  | **Dysmenorrhea** | | **HMB** | |
| --- | --- | --- | --- | --- |
|  | **Cases**  **n (%)**  (unless otherwise specified) | **Controls**  **n (%)**  (unless otherwise specified) | **Cases**  **n (%)**  (unless otherwise specified) | **Controls**  **n (%)**  (unless otherwise specified) |
|  | **641** | **1,254** | **527** | **2,083** |
| **Prenatal** | | | | |
| Maternal university degree* | 70 (10.9) | 208 (16.6) | 46 (8.7) | 361 (17.3) |
| Missing | 72 (11.2) | 137 (10.9) | 65 (12.3) | 188 (9.0) |
| Alcohol exposure during pregnancy | 394 (61.5) | 785 (62.6) | 317 (60.2) | 1,336 (64.1) |
| Missing | 151 (23.6) | 258 (20.6) | 129 (24.5) | 397 (19.1) |
| Smoke exposure during pregnancy | 142 (22.2) | 222 (17.7) | 132 (25.0) | 364 (17.5) |
| Missing | 95 (14.8) | 184 (14.7) | 86 (16.3) | 282 (13.5) |
| Any HDP | 113 (17.6) | 201 (16.0) | 97 (18.4) | 310 (14.9) |
| Missing | 44 (6.9) | 67 (5.3) | 41 (7.8) | 104 (5.0) |
| Preeclampsia | 9 (1.4) | 26 (2.1) | 11 (2.1) | 42 (2.0) |
| Missing | 44 (6.9) | 67 (5.3) | 41 (7.8) | 104 (5.0) |
| **Age 13 years** | | | | |
| Have drunk alcohol before | 165 (25.7) | 242 (19.3) | 137 (26.0) | 430 (20.6) |
| Missing | 311 (48.5) | 649 (51.8) | 240 (45.5) | 1,013 (48.6) |
| Have smoked cigarettes before* | 99 (15.4) | 92 (7.3) | 91 (17.3) | 160 (7.7) |
| Missing | 160 (25.0) | 383 (30.5) | 130 (24.7) | 544 (26.1) |
| **End of puberty** |  |  |  |  |
| Contraception use* | 445 (69.4) | 271 (21.6) | 390 (74.0) | 511 (24.5) |
| Missing | 3 (0.5) | 7 (0.6) | 2 (0.004) | 11 (0.005) |
| Comorbidity reported* | 39 (6.1) | 30 (2.4) | 36 (6.8) | 58 (2.8) |
| Missing | 363 (56.6) | 837 (66.7) | 301 (57.1) | 1292 (62.0) |

*Identified characteristic *a priori*.

**Supplementary Table 2** Mean (SD) of each continuous characteristic in cases and controls for each symptom identified in the hypothesis-generating EWAS and missing data in each variable

|  | **Dysmenorrhea** | | | | **HMB** | | | |
| --- | --- | --- | --- | --- | --- | --- | --- | --- |
|  | **Cases**  **n (%)**  (unless otherwise specified) | | **Controls**  **n (%)**  (unless otherwise specified) | | **Cases**  **n (%)**  (unless otherwise specified) | **Controls**  **n (%)**  (unless otherwise specified) | | |
|  | **641** | | | **1,254** | **527** | | **2,083** | |
| **Prenatal** | | | | | | | | |
| Mean maternal BMI, kg/m^2^ (SD) | 23.1 (3.9) | | 22.9 (3.9) | | 23.1 (4.0) | 22.8 (3.7) | | |
| Missing | 94 (14.7) | | 201 (16.0) | | 80 (15.1) | 301 (14.4) | | |
| **Delivery** | | | | | | | | |
| Mean gestational age (SD) | 39.6 (1.7) | | 39.4 (1.8) | | 39.5 (1.8) | 39.6 (1.8) | | |
| Missing | 39 (6.1) | | 64 (5.1) | | 35 (6.6) | 100 (4.8) | | |
| **Age 7 years** | | | | | | | | |
| Mean BMI, kg/m^2^ (SD) | 16.8 (2.6) | | 16.3 (2.1) | | 16.8 (2.5) | 16.3 (2.1) | | |
| Missing | 137 (21.4) | | 318 (25.4) | | 114 (21.6) | 450 (21.6) | | |
| Mean cholesterol, mmol/L | 4.5 (0.7) | | 4.5 (0.7) | | 4.5 (0.7) | 4.5 (0.7) | | |
| Missing | 309 (48.2) | | 636 (50.7) | | 253 (48.0) | 1,008 (48.4) | | |
| Mean cotinine, ng/mL (SD) | 1.5 (1.4) | | 1.2 (1.2) | | 1.5 (1.5) | 1.2 (1.1) | | |
| Missing | 302 (48.2) | | 617 (49.2) | | 254 (48.2) | 978 (47.0) | | |
| **Age 8 years** | | | | | | | | |
| Mean non-word repetition score, (SD) | | 7.2 (2.5) | | 7.3 (2.6) | 7.1 (2.4) | | | 7.4 (2.5) |
| Missing | | 62 (9.7) | | 368 (29.3) | 145 (27.5) | | | 527 (25.3) |
| **Age 9 years** | | | | | | | | |
| Mean C-reactive protein, mmol/L (SD) | | 1.2 (3.7) | | 0.7 (1.4) | 1.2 (3.2) | | | 0.8 (2.7) |
| Missing | | 322 (50.2) | | 625 (49.8) | 279 (52.9) | | | 981 (47.1) |
| **Puberty** | | | | | | | | |
| Mean age, months (SD)* | 146.0 (12.8) | | 151.5 (12.6) | | 146.4 (13.3) | 150.4 (12.3) | | |
| Missing | 123 (19.2) | | 331 (26.4) | | 91 (17.3) | 478 (22.9) | | |
| **Age 16 years** | | | | | | | | |
| Mean ACE score (SD) | 2.0 (1.5) | | 1.6 (1.7) | | 2.1 (1.9) | 1.5 (1.5) | | |
| Missing | 419 (65.4) | | 892 (71.1) | | 362 (68.7) | 1,433 (68.8) | | |

**Supplementary Table 3** Comparison of dysmenorrhea cases and controls for each binary characteristic (as shown in Figure 3)

|  | **OR**  **(95% CI)** | ***p*-value** | **aOR**  **(SEP only)**  **(95% CI)** | ***p*-value** | **aOR**  **(AAM & SEP)** | **(95% CI)** |
| --- | --- | --- | --- | --- | --- | --- |
| **Prenatal** |  |  |  |  |  |  |
| Maternal university degree* | 0.61  (0.46 to 0.82) | 1.02$\times{10}^{-3}$ | - | - | - | - |
| Alcohol exposure during pregnancy | 1.10  (0.84 to 1.45) | 0.476 | 1.16  (0.88 to 1.52) | 0.303 | 1.14  (0.83 to 1.57) | 0.403 |
| Smoke exposure during pregnancy | 1.34  (1.05 to 1.71) | 1.69$\times{10}^{-2}$ | 1.22  (0.94 to 1.59) | 0.129 | 1.09  (0.81 to 1.48) | 0.571 |
| Any HDP | 1.15  (0.89 to 1.48) | 0.297 | 1.15  (0.89 to 1.50) | 0.290 | 1.02  (0.75 to 1.38) | 0.919 |
| Preeclampsia | 0.68  (0.32 to 1.47) | 0.329 | 0.73  (0.34 to 1.59) | 0.428 | 0.66  (0.28 to 1.60) | 0.362 |
| **Age 13 years** |  |  |  |  |  |  |
| Have drunk alcohol before | 1.54  (1.18 to 2.02) | 1.67$\times{10}^{-3}$ | 1.51  (1.13 to 2.00) | 4.64$\times{10}^{-3}$ | 1.37  (0.99 to 1.89) | 5.61$\times{10}^{-2}$ |
| Have smoked cigarettes before* | 2.19  (1.61 to 2.99) | 6.25$\times{10}^{-7}$ | 2.23  (1.60 to 3.12) | 2.34$\times{10}^{-6}$ | 1.61  (1.11 to 2.33) | 1.23$\times{10}^{-2}$ |
| **End of puberty** |  |  |  |  |  |  |
| Contraception use* | 8.30  (6.70 to 10.3) | 3.13$\times{10}^{-82}$ | 7.76  (6.18 to 9.76) | 3.81$\times{10}^{-69}$ | 7.55  (5.80 to 9.82) | 2.34$\times{10}^{-51}$ |
| Comorbidity reported* | 2.11  (1.27 to 3.48) | 3.70$\times{10}^{-5}$ | 1.60  (0.93 to 2.76) | 0.089 | 1.76  (0.92 to 3.37) | 0.086 |

**Supplementary Table 4** Comparison of dysmenorrhea cases and controls for each continuous characteristic (as shown in Figure 4)

| **Dysmenorrhea cases vs controls** | **OR**  **(95% CI)** | ***p*-value** | **aOR**  **(SEP only)**  **(95% CI)** | ***p*-value** | **aOR**  **(AAM & SEP)**  **(95% CI)** | ***p*-value** |
| --- | --- | --- | --- | --- | --- | --- |
| **Prenatal** | | | | | | |
| Maternal BMI | 1.05  (0.94 to 1.16) | 0.390 | 1.02  (0.92 to 1.14) | 0.682 | 0.95  (0.84 to 1.07) | 0.421 |
| **Delivery** | | | | | | |
| Gestational age | 1.35  (0.98 to 1.86) | 0.065 | 1.32  (0.95 to 1.84) | 0.094 | 1.41  (0.97 to 2.06) | 0.074 |
| **Age 7 years** | | | | | | |
| BMI | 1.20  (1.09 to 1.33) | 2.05$\times{10}^{-4}$ | 1.18  (1.06 to 1.31) | 2.82$\times{10}^{-3}$ | 0.99  (0.88 to 1.13) | 0.92 |
| Cholesterol | 0.98  (0.86 to 1.11) | 0.72 | 0.97  (0.85 to 1.12) | 0.72 | 0.88  (0.75 to 1.03) | 0.12 |
| Cotinine | 1.22  (1.07 to 1.41) | 3.93$\times{10}^{-3}$ | 1.15  (0.99 to 1.33) | 7.43$\times{10}^{-2}$ | 1.16  (0.97 to 1.38) | 0.11 |
| **Age 8 years** |  |  |  |  |  |  |
| Non-word repetition | 0.95  (0.85 to 1.06) | 0.34 | 0.98  (0.87 to 1.11) | 0.75 | 0.95  (0.83 to 1.08) | 0.42 |
| **Age 9 years** |  |  |  |  |  |  |
| CRP | 1.29  (1.07 to 1.54) | 6.09$\times{10}^{-3}$ | 1.30  (1.06 to 1.60) | 1.24$\times{10}^{-2}$ | 1.18  (0.95 to 1.47) | 0.13 |
| **Teen** | | | | | | |
| Age at menarche* | 0.66  (0.59 to 0.73) | 4.60$\times{10}^{-14}$ | 0.64  (0.57 to 0.72) | 1.02$\times{10}^{-13}$ | - | - |
| **Age 16 years** | | | | | | |
| ACE score | 1.33  (1.15 to 1.53) | 8.52$\times{10}^{-5}$ | 1.33  (1.15 to 1.53) | 1.08$\times{10}^{-4}$ | 1.30  (1.11 to 1.53) | 1.17$\times{10}^{-3}$ |

**Supplementary Table 5** Comparison of HMB cases and controls for each binary characteristic (as shown in Figure 3)

| **Dysmenorrhea cases vs controls** | **OR**  **(95% CI)** | ***p*-value** | **aOR**  **(SEP only)**  **(95% CI)** | ***p*-value** | **aOR**  **(AAM & SEP)** | **(95% CI)** |
| --- | --- | --- | --- | --- | --- | --- |
| **Prenatal** |  |  |  |  |  |  |
| Maternal university degree* | 0.47  (0.34 to 0.65) | 5.38$\times{10}^{-6}$ | - | - | - | - |
| Alcohol exposure during pregnancy | 1.03  (0.78 to 1.34) | 0.857 | 1.10  (0.84 to 1.45) | 0.487 | 1.12  (0.82 to 1.52) | 0.472 |
| Smoke exposure during pregnancy | 1.69  (1.33 to 2.13) | 1.20$\times{10}^{-5}$ | 1.50  (1.17 to 1.93) | 1.47$\times{10}^{-3}$ | 1.34  (1.01 to 1.79) | 4.32$\times{10}^{-2}$ |
| Any HDP | 1.34  (1.04 to 1.73) | 2.27$\times{10}^{-2}$ | 1.35  (1.04 to 1.75) | 2.61$\times{10}^{-2}$ | 1.19  (0.89 to 1.61) | 0.242 |
| Preeclampsia | 1.07  (0.55 to 2.09) | 0.848 | 1.10  (0.56 to 2.17) | 0.784 | 1.03  (0.48 to 2.20) | 0.928 |
| **Age 13 years** |  |  |  |  |  |  |
| Have drunk alcohol before | 1.36  (1.05 to 1.77) | 2.16$\times{10}^{-2}$ | 1.37  (1.04 to 1.80) | $2.74\times{10}^{-2}$ | 1.35  (1.00 to 1.82) | 4.67$\times{10}^{-2}$ |
| Have smoked cigarettes before* | 2.56  (1.93 to 3.14) | 1.05$\times{10}^{-10}$ | 2.75  (2.03 to 3.73) | $7.56\times{10}^{-11}$ | 2.35  (1.69 to 3.26) | 3.52$\times{10}^{-7}$ |
| **End of puberty** |  |  |  |  |  |  |
| Contraception use* | 8.82  (7.08 to 11.0) | 4.90$\times{10}^{-84}$ | 9.19  (7.24 to 11.7) | 2.55$\times{10}^{-74}$ | 9.77  (7.46 to 12.8) | 9.28$\times{10}^{-62}$ |
| Comorbidity reported* | 2.39  (1.53 to 3.74) | 1.22$\times{10}^{-4}$ | 2.66  (1.64 to 4.31) | 7.39$\times{10}^{-6}$ | 2.71  (1.56 to 4.70) | 3.88$\times{10}^{-5}$ |

**Supplementary Table 6** Comparison of HMB cases and controls for each continuous characteristic (as shown in Figure 4)

| **HMB case vs control** | **OR**  **(95% CI)** | ***p*-value** | **aOR**  **(SEP only)**  **(95% CI)** | ***p*-value** | **aOR**  **(AAM & SEP)**  **(95% CI)** | ***p*-value** |
| --- | --- | --- | --- | --- | --- | --- |
| **Prenatal** | | | | | | |
| Maternal BMI | 1.08  (0.97 to 1.19) | 0.164 | 1.05  (0.94 to 1.17) | 0.391 | 0.99  (0.88 to 1.12) | 0.922 |
| **Delivery** | | | | | | |
| Gestational age | 0.91  (0.67 to 1.22) | 0.520 | 0.91  (0.66 to 1.26) | 0.575 | 0.92  (0.64 to 1.33) | 0.671 |
| **Age 7 years** | | | | | | |
| BMI | 1.24  (1.13 to 1.37) | 1.52$\times{10}^{-5}$ | 1.22  (1.09 to 1.35) | 2.68$\times{10}^{-4}$ | 1.12  (0.99 to 1.26) | 7.43$\times{10}^{-2}$ |
| Cholesterol | 1.05  (0.92 to 1.19) | 0.49 | 1.00  (0.87 to 1.15) | 0.99 | 1.00  (0.86 to 1.16) | 0.98 |
| Cotinine | 1.25  (1.09 to 1.43) | 1.09$\times{10}^{-3}$ | 1.18  (1.01 to 1.36) | 3.37$\times{10}^{-2}$ | 1.18  (1.00 to 1.40) | 5.70$\times{10}^{-2}$ |
| **Age 8 years** |  |  |  |  |  |  |
| Non-word repetition | 0.90  (0.80 to 1.00) | 5.18$\times{10}^{-2}$ | 0.96  (0.85 to 1.08) | 0.50 | 0.93  (0.81 to 1.06) | 0.27 |
| **Age 9 years** |  |  |  |  |  |  |
| CRP | 1.12  (1.00 to 1.27) | 5.45$\times{10}^{-2}$ | 1.10  (0.97 to 1.24) | 0.13 | 1.04  (0.92 to 1.17) | 0.54 |
| **Puberty** | | | | | | |
| Age at menarche* | 0.73  (0.66 to 0.81) | 4.17$\times{10}^{-9}$ | 0.73  (0.65 to 0.81) | 2.08$\times{10}^{-8}$ | - | - |
| **Age 16 years** | | | | | | |
| ACE score | 1.36  (1.18 to 1.56) | 1.30$\times{10}^{-5}$ | 1.36  (1.18 to 1.57) | 2.09$\times{10}^{-5}$ | 1.35  (1.15 to 1.57) | 1.51$\times{10}^{-4}$ |

##### Sensitivity analysis

**Supplementary Table 7** Differentially methylated CpG sites in hypothesis-generating EWAS with comorbidity cases removed compared with the effect estimates from the primary EWAS (dysmenorrhea)

| **Probe ID** | $\boldsymbol{\beta}$  (ex. comorbidities)  (95%CI) | ***p*-value** | $\boldsymbol{\beta}$  (primary analysis)  (95%CI) | ***p*-value** | **Absolute % change** |
| --- | --- | --- | --- | --- | --- |
| cg08142094 | -0.038  (-0.053 to -0.023) | 2.73${\times10}^{-6}$ | -0.036  (-0.050 to -0.022) | 1.02${\times10}^{-6}$ | +5% |
| cg23012731 | -0.009  (-0.013 to -0.006) | 5.12${\times10}^{-7}$ | -0.008  (-0.011 to -0.005) | 3.53${\times10}^{-6}$ | +12.5% |
| cg04737758 | 0.036  (0.018 to 0.054) | 8.97${\times10}^{-5}$ | 0.042  (0.025 to 0.058) | 1.13${\times10}^{-6}$ | -14.3% |
| cg15017604 | 0.030  (0.017 to 0.043) | 8.87${\times10}^{-6}$ | 0.026  (0.015 to 0.037) | 9.39${\times10}^{-6}$ | +15.4% |

**Supplementary Table 8** Differentially methylated CpG sites in hypothesis-generating EWAS with comorbidity cases removed compared with the effect estimates from the primary EWAS (HMB)

| **Probe ID** | $\boldsymbol{\beta}$  (ex. comorbidities)  (95%CI) | ***p*-value** | $\boldsymbol{\beta}$  (primary analysis)  (95%CI) | ***p*-value** | **Absolute % change** |
| --- | --- | --- | --- | --- | --- |
| cg24196053 | -0.011  (-0.016 to -0.005) | 8.33${\times10}^{-5}$ | -0.010  (-0.014 to -0.006) | 1.50${\times10}^{-5}$ | +10% |

**Supplementary Figure 1** Coefficient plot representing binary characteristics associated with dysmenorrhea and HMB with participants with a comorbidity removed

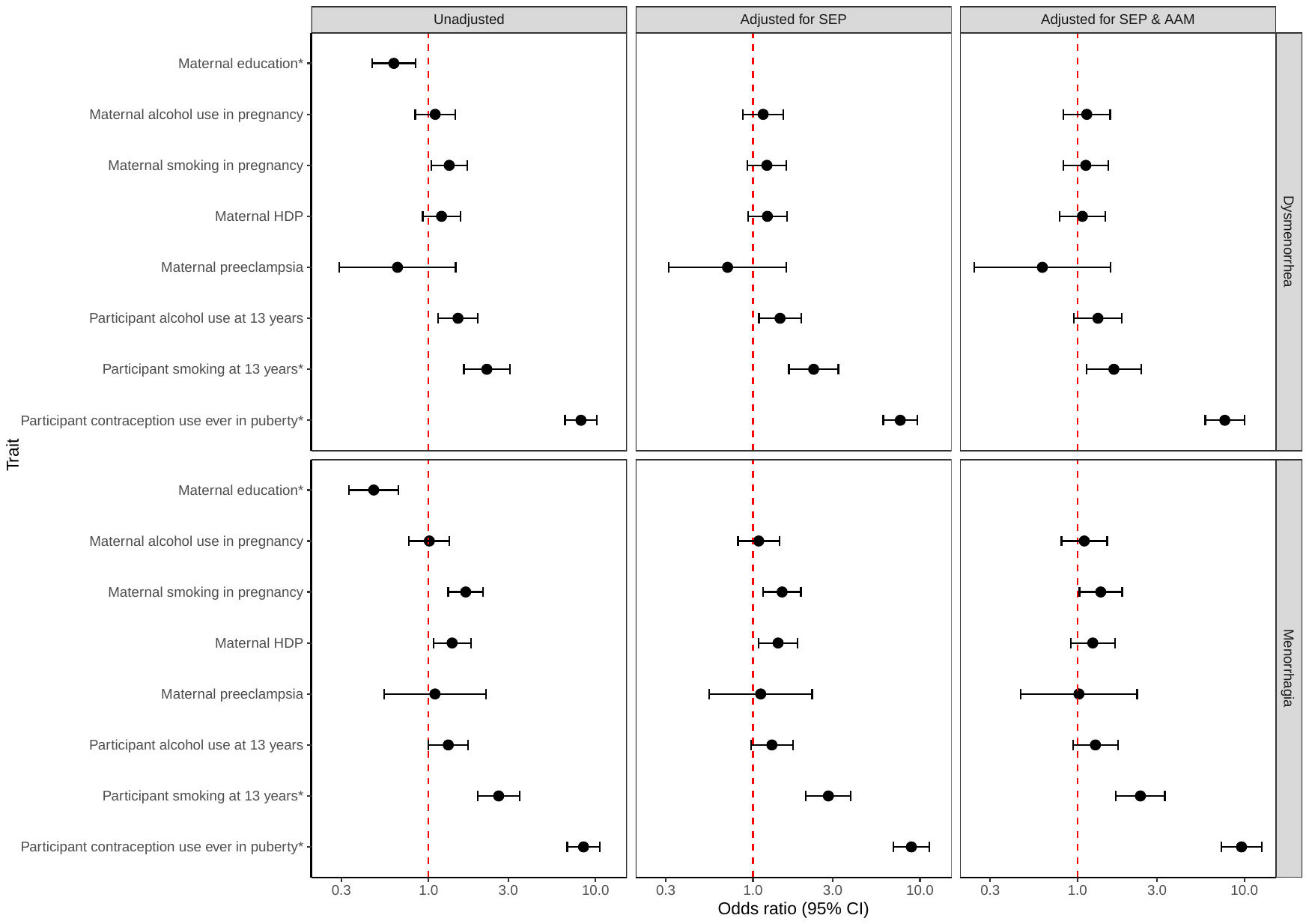

* Identified as an associated characteristic *a priori*

**Supplementary Figure 2** Coefficient plot representing continuous characteristics associated with dysmenorrhea and HMB with participants with a comorbidity removed

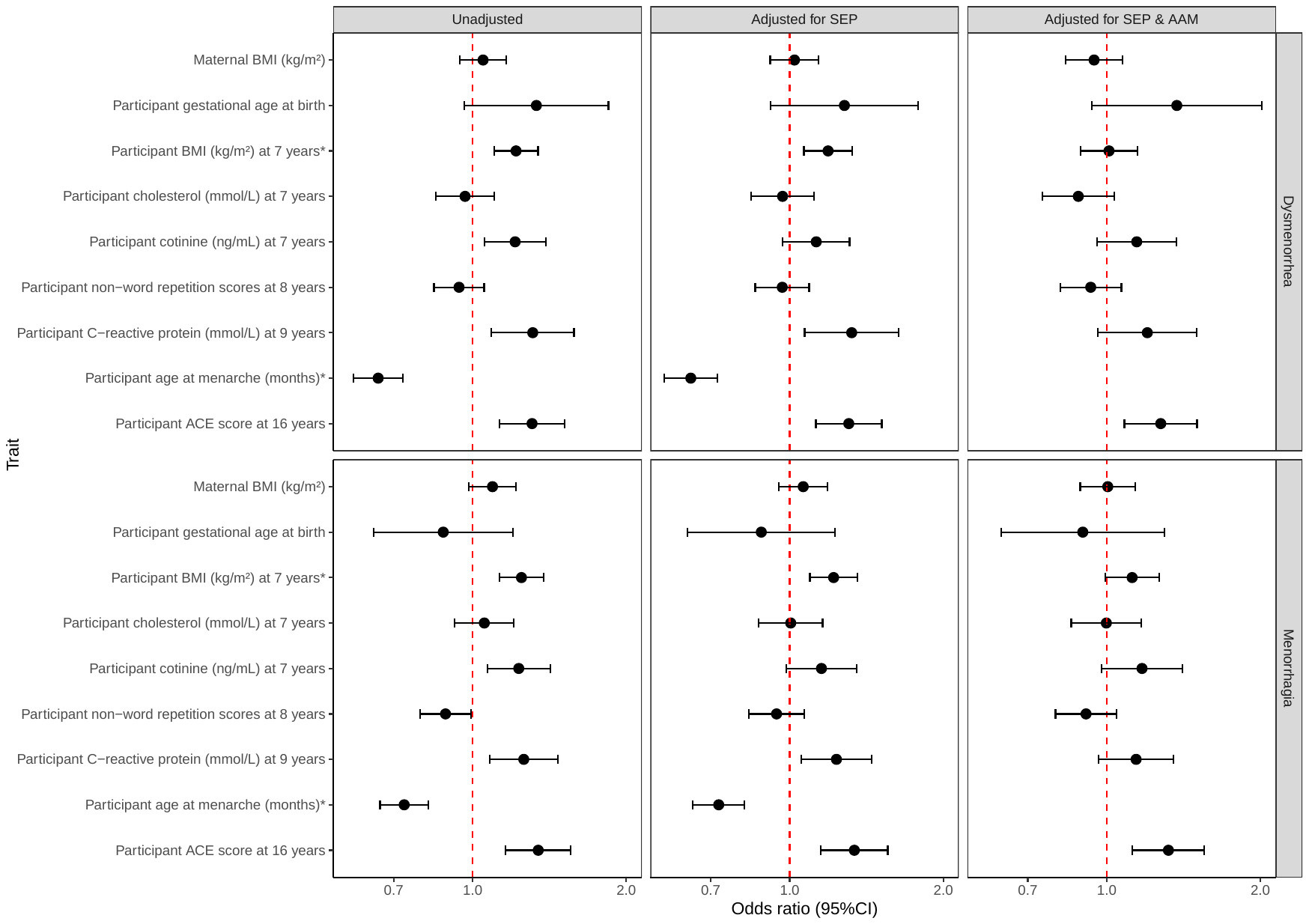

* Identified as an associated characteristic *a priori*

**Supplementary Figure 3** Coefficient plot representing binary traits associated with less severe dysmenorrhea and HMB

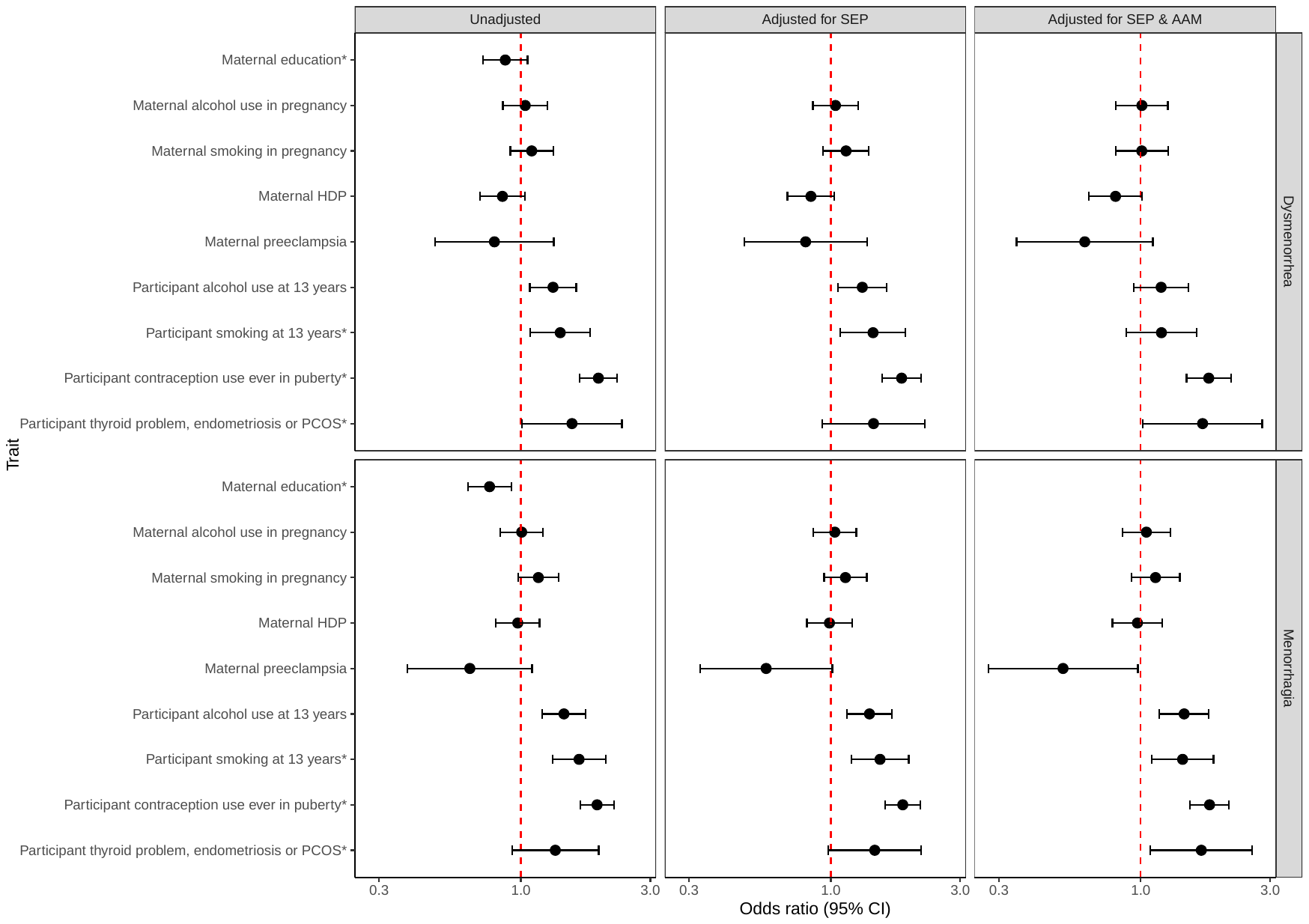

* Identified as an associated trait *a priori*

**Supplementary Figure 4** Coefficient plot representing continuous traits associated with less severe dysmenorrhea and HMB

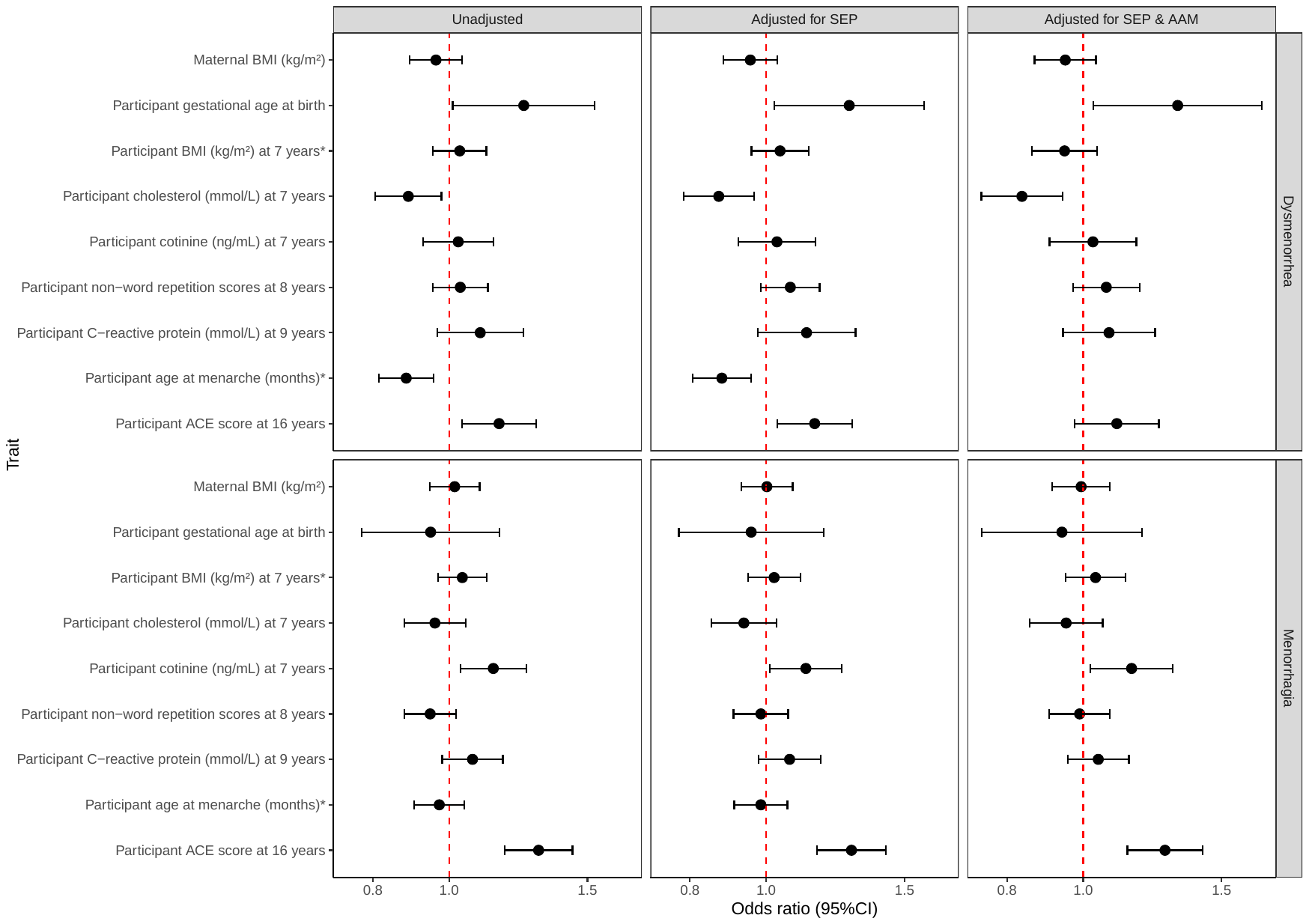

* Identified as an associated trait *a priori*
